# Disruption of central dopamine metabolism in infants with severe spinal muscular atrophy

**DOI:** 10.64898/2026.02.28.26347004

**Authors:** Tommaso Nuzzo, Barbara Risi, Valentina Bassareo, Adele D’Amico, Alberto Imarisio, Antonella Longo, Manolo Carta, Chiara Panicucci, Claudio Bruno, Enza Maria Valente, Massimiliano Filosto, Enrico Bertini, Francesco Errico, Alessandro Usiello

## Abstract

Spinal muscular atrophy (SMA) is a severe neuromuscular disorder caused by reduced expression of the survival motor neuron (SMN) protein. In addition to affecting motor neuron survival, SMN deficiency impacts multisystem physiology and neurotransmission. Dopaminergic dysfunction has been reported in mouse models of SMA, leading to postural and locomotor impairments that improve upon treatment with L-DOPA and benserazide. However, whether altered dopamine metabolism contributes to clinical symptoms in SMA patients remains unclear. To investigate this issue, we conducted a real-world observational study involving pediatric patients with SMA1, SMA2, and SMA3. We performed a longitudinal measurement of the main dopamine-related catabolites – 3,4-dihydroxyphenylacetic acid (DOPAC) and homovanillic acid (HVA) – in cerebrospinal fluid (CSF) samples collected at baseline and after five intrathecal doses of Nusinersen, an SMN-enhancing therapy. No significant differences were observed in CSF DOPAC and HVA levels across SMA types or following treatment, and no association emerged with SMN2 copy number. In contrast, lower baseline DOPAC levels were detected in SMA1 patients requiring gastrostomy and tracheostomy, and were associated with reduced improvement on the CHOP-INTEND scale. These findings suggest that reduced central dopaminergic turnover reflects disease progression in SMA1 and is associated with more severe clinical impairment and limited functional recovery.

## Introduction

Spinal muscular atrophy (SMA) is a rare, inherited disease characterized by degeneration of lower motor neurons^1,2^, due to homozygous deletions or mutations in the *survival motor neuron 1* (*SMN1)* gene^3^, resulting in reduced levels of SMN protein. In the last decade, disease-modifying therapies targeting the alternative splicing of the paralogous gene *SMN2* have profoundly altered the natural history of SMA. These include Nusinersen (Spinraza), an antisense oligonucleotide administered intrathecally every 4 months (maintenance regimen)^4^, Risdiplam (Evrysdi), a small molecule administered orally on a daily basis^5^ and, more recently, gene replacement therapy using adeno-associated viral vectors^6,7^. Irrespective of the specific mechanism of action, these therapeutic approaches increase SMN protein levels and have significantly improved survival and motor outcomes in pediatric patients, particularly SMA1, which was uniformly fatal in the pre-treatment era^8^.

Despite these advances, the clinical benefit of SMN-enhancing therapies remains incomplete, especially when treatment is delayed or in patients with a long disease history, probably due to the irreversible loss of motor neuron pools associated with disease progression^9,10^. More generally, neither therapy has proved capable of completely correcting the symptoms. The incomplete knowledge of the pathogenic effects of SMN deficiency across the central nervous system (CNS) and peripheral organs, along with the absence of reliable biomarkers reflecting the clinical response to SMN induction, represent current limitations to understanding why the current therapies are only partially effective in treating SMA.

SMN is expressed ubiquitously and plays a key role in multiple cellular processes^11^, including a well-characterized housekeeping function in ribonucleoprotein assembly^12^. Due to its diverse functions, a deficiency in SMN has been linked to significant metabolic dysregulation, impacting the balance of lipids, glucose, and amino acids in both SMA models and patients^13–24^. In particular, recent OMIC-based studies have shown that Nusinersen significantly alters the central metabolism of several amino acids, including L-glutamate, L-glutamine, L-serine, L-arginine, branched amino acids (BrAAs), and the aromatic amino acids^23,25–28^. While the mechanistic link between SMN deficiency and amino acid dysregulation remains to be fully elucidated, these findings point to a potential disruption of neurotransmission in SMA. Beyond their primary metabolic roles in protein synthesis and energy homeostasis, these amino acids are directly involved in synaptic transmission as neurotransmitters or serve as their precursors. Accordingly, a growing body of preclinical evidence indicates that synaptic dysfunction represents a central pathogenic feature of SMA, affecting both excitatory and inhibitory circuits within the spinal cord and higher-order CNS networks^25–27,29–32^.

Although much of the earlier work has focused on glutamatergic neurotransmission dysfunction at sensory-motor synapses, increasing attention has recently been directed toward monoaminergic systems. In this context, SMN deficiency has been shown to alter dopamine, norepinephrine and serotonin metabolism in the brain and spinal cord of SMNΔ7 mice, a widely used and well-characterized transgenic model of severe SMA^33^, especially during the later stages of the disease^28^. In line with these observations, clinical data indicate that untreated SMA1 patients exhibit elevated cerebrospinal fluid (CSF) levels of aromatic amino acids compared to healthy controls, and that Nusinersen treatment increased norepinephrine levels^28^. Additionally, a recent *in vivo* study showed that loss of dopaminergic synapses in vulnerable spinal motor neurons causes locomotor and postural deficits in the SMNΔ7 mouse model, and that these deficits were partially rescued by pharmacological enhancement of dopaminergic signaling using oral L-DOPA and benserazide administration^34^. Together, earlier results suggest that alterations in neurotransmitter signaling in SMA affect not only spinal motor neuron circuitry but also descending projections from monoaminergic pathways^29–32,34–36^.

Based on these emerging findings linking SMN deficiency to abnormalities of catecholaminergic neurotransmission, in the present work we investigated central dopamine metabolism in SMA patients in a real-world setting. We quantified the CSF levels of the primary dopamine-related metabolites - 3,4-dihydroxyphenylacetic acid (DOPAC) and homovanillic acid (HVA) - in drug-free SMA patients of varying disease severity. As dopamine is rapidly unstable and prone to rapid oxidation, DOPAC and HVA represent reliable surrogate markers of dopamine metabolism in the CSF^37,38^. Further, we investigated the impact of Nusinersen on these metabolites in treated patients and explored their relationship with motor outcomes, in light of recent preclinical evidence linking dopaminergic dysfunction to SMA motor impairment^34^.

## Methods

### Patients

This was a real-world, longitudinal two-center cohort study conducted on 55 patients affected by SMA1 (n=26), SMA2 (n=17), and SMA3 (n=12), who received intrathecal treatment with Nusinersen (12 mg) at the Bambino Gesù Hospital (Rome, Italy) and at the Giannina Gaslini Institute (Genoa, Italy). The study was approved by the local Ethics Committee of the two Hospitals (2395_OPBG_2021). Written informed consent was obtained from all participants and/or their legal guardians. All relevant ethical regulations for human research participants were followed.

All patients were clinically and genetically diagnosed, and the number of copies of the *SMN2* gene was determined. All SMA1 patients, regardless of age or disease severity, participated in the Expanded Access Program (EAP) for compassionate use to patients with the infantile form only, which occurred in Italy between November 2016 and November 2017. The overall clinical response to Nusinersen treatment in these patients^39,40^ as well as in SMA2 and SMA3 patients ^41^ has previously been reported as part of the full Italian cohort.

Demographic and clinical features of patients are reported in Table 1. CSF samples and clinical data collected as per standard protocol at day 0 (T0; baseline) and day 302 (T302; after 5 doses of Nusinersen) (from a subgroup of 44 patients) were evaluated for this study. In addition, CSF samples from pediatric control subjects without neurological disorders, aged 7.5–11.5 years (n=3), were used as a reference for physiological CSF baseline levels of DOPAC and HVA. These samples were not included in statistical analyses due to the small sample size.

**Table 1.**
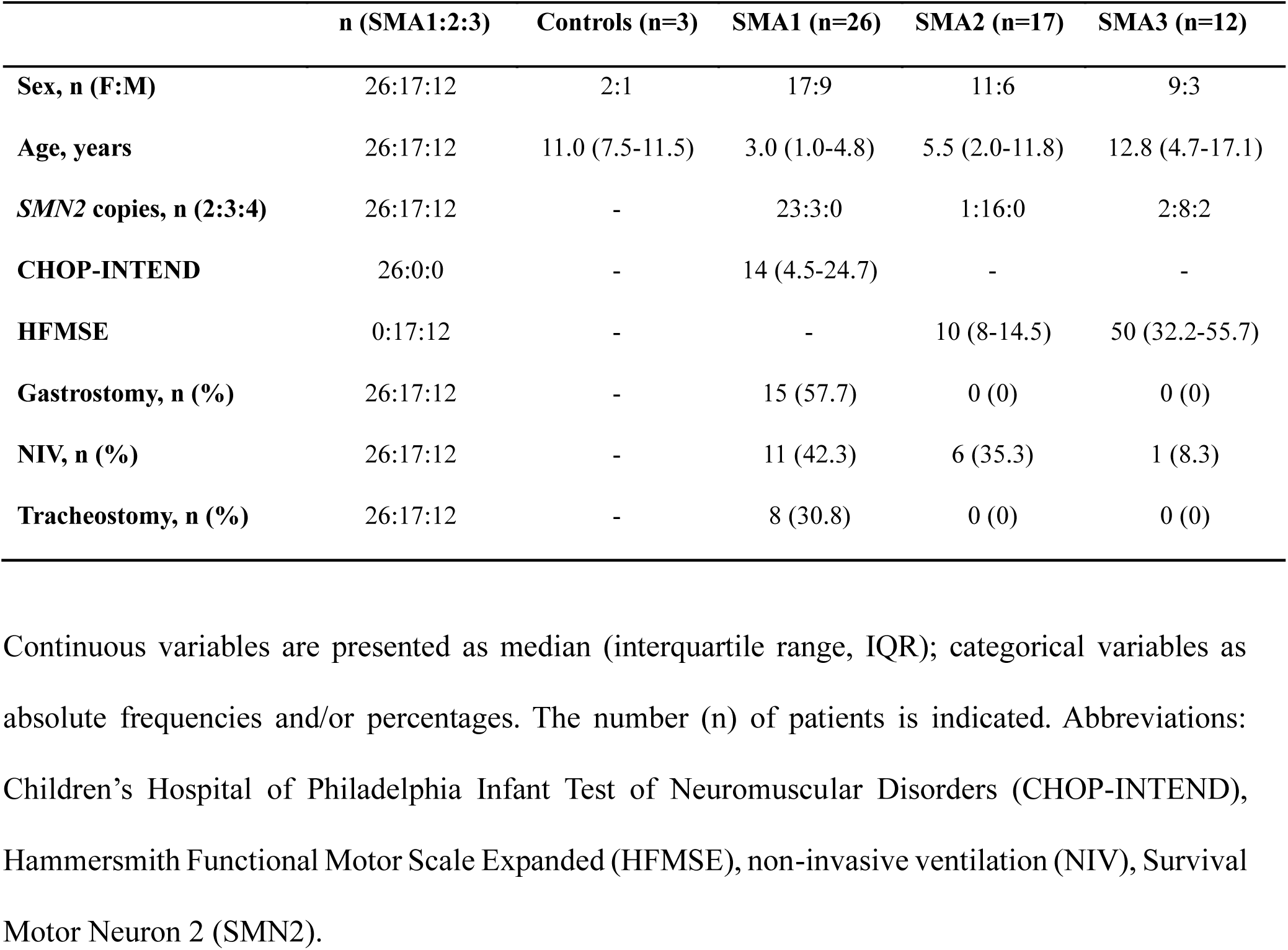
Demographic and clinical characteristics of treatment-naïve SMA1, SMA2 and SMA3 patients and healthy controls enrolled in the study.

### Clinical assessments

Patients were assessed at baseline (T0) and after the fifth infusion of Nusinersen (T302) using the CHOP-INTEND (Children’s Hospital of Philadelphia Infant Test of Neuromuscular Disorders)^42,43^ or HFMSE (Hammersmith Functional Motor Scale Expanded)^44,45^, based on patients’ functional abilities and age. Ventilation requirement, either non-invasive (NIV) or invasive (tracheostomy) and feeding support (gastrostomy) were also collected (Table 1).

At baseline, all SMA1 patients were non-sitters, with CHOP-INTEND scores ranging from 4.5 to 24.7. HFMSE scores ranged from 8 to 14.5 in SMA2 patients and from 32.2 to 55.7 in SMA3 patients. At the start of the treatment, 20 of the 26 SMA1 patients were older than 1 year, with ages ranging from 1 year and 1 month to 7 years and 8 months. Eight patients had tracheostomy, eleven were under non-invasive ventilation (NIV) for less than 16 h/day and seven patients were in spontaneous breathing. Fifteen patients had gastrostomy. The age of the seventeen SMA2 patients ranged from 11 months to 13 years and 3 months at baseline. Six of these patients were under non-invasive ventilation (NIV) and none had gastrostomy or tracheostomy. Among the twelve SMA3 patients, one was under NIV for less than 16 h/day and none had gastrostomy.

### Intrathecal treatment with Nusinersen and CSF collection

Intrathecal administration of Nusinersen (12 mg) was performed in a hospital setting, as previously reported^23^. CSF samples were collected in polypropylene tubes and stored at −80°C until further analysis. Among the 26 SMA1 patients, CSF samples were obtained from all patients at first injection (T0), and from 17 patients at T302. Among the 17 SMA2 patients, CSF samples were collected from all patients at T0 and from 15 patients at T302. For the 12 SMA3 patients, CSF samples were collected from all patients at both T0 and T302. The sample size was determined by the number of eligible SMA patients treated at the two centers during the study period; no formal sample size calculation was performed due to the exploratory nature of the study. Missing follow-up CSF samples were due to clinical or procedural reasons; no imputation was performed. Loss to follow-up was handled by restricting longitudinal analyses to patients with paired T0–T302 samples. Exclusion criteria included the presence of symptoms or laboratory abnormalities in blood biochemical and haematological parameters suggestive of a systemic inflammatory state, as well as the use of immunosuppressive treatments within the 6 months preceding enrollment.

### HPLC detection

Twenty μl samples (5 μl of hydrophilic phase from CSF + 15 μl of 0.2 N PCA) were injected without purification into an HPLC equipped with a reverse phase column (LC-18 DB, 15 cm, 5 μm particle size, Supelco, Waters, Milford, MA, USA) and a coulometric detector (ESA, Coulochem II, Bedford, MA, USA) to quantify DOPAC and HVA.

To detect these catabolites, the first electrode of the detector was set at +125 mV (oxidation) and the second at −175 mV (reduction). The composition of the mobile phase was (in mM): 50 NaH2PO4, 0.1 Na2-EDTA, 0.5 n-octyl sodium sulfate, 15% (v/v) methanol, pH 3.793,94. The composition of the mobile phase was: 120 mM CH3-COONa, 100 mM citric acid, 0.3 mM EDTA, 5% (v/v) methanol, pH 4.995. For quantitative determination of each neurotransmitter, calibration curves were run using standards (SIGMA–ALDRICH, MO, USA). For data acquisition, the software ESA CDS (Euroservice, Ge, Italy) was used. Final values were expressed as nM. All samples were analyzed using the same instrumentation, analytical conditions, and calibration procedures to minimize measurement bias.

### Statistical Analysis

Statistical analyses were performed using SPSS software v.26 (SPSS Inc., Chicago, IL, USA), with the statistical significance level set to *p*≤0.05. Normality distribution was assessed by the Kolmogorov–Smirnov and Shapiro-Wilk tests. Continuous variables were summarized as the median and interquartile range (IQR), while qualitative variables by absolute or relative frequency. Differences between SMA groups were studied by non-parametric Kruskal-Wallis test followed, if statistically significant, by post-hoc tests performed by Mann-Whitney test with Bonferroni’s correction. The effect of confounders (sex, age) was evaluated by ANCOVA on natural log-transformed variables. ANCOVA, corrected for sex and age, was also applied to test for differences between patients with and without gastrostomy or tracheostomy. Differences between dependent groups (T0-T302) were studied by Wilcoxon Matched Pairs Signed Ranks Test. Association between continuous variables was evaluated by Spearman’s correlation, corrected by age as a confounder. Continuous variables were analyzed as raw or log-transformed values depending on distributional assumptions. No sensitivity analyses were performed due to the exploratory nature of the study.

## Results

### Analysis of dopamine-related metabolites in the CSF of SMA patients at baseline and after Nusinersen treatment

We investigated whether SMN deficiency affects the concentrations of dopamine-related metabolites, measuring DOPAC and HVA by HPLC (Figure 1a) in the CSF of treatment-naïve SMA1 (n=26), SMA2 (n=17) and SMA3 (n=12) patients, whose clinical and demographic features are detailed in Table 1. Non-parametric Kruskal-Wallis analysis showed comparable CSF concentrations of both DOPAC and HVA among naïve SMA1, SMA2, and SMA3 patients (Figure 1b,c, Suppl. Table 1). We also confirmed these findings by age- and sex-adjusted ANCOVA analysis (Suppl. Table 1). Consistent with previous reports in pediatric reference populations and metabolic disease settings^46–48^, we observed a negative correlation between DOPAC levels and age in SMA1 patients (r=-0.451, p=0.021; Suppl. Table 2).

**Figure 1.**
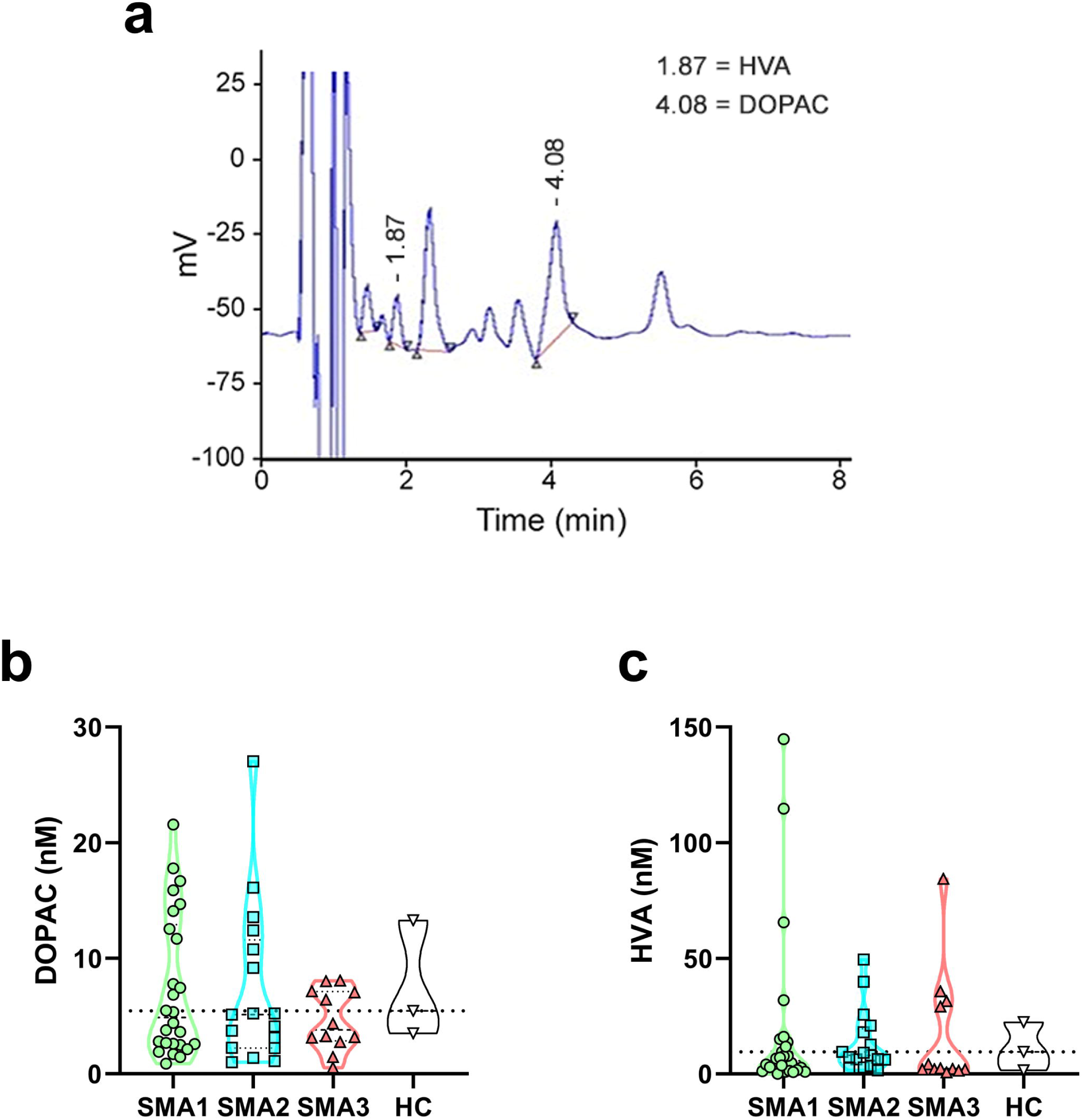
CSF DOPAC and HVA levels in SMA patients and control individuals. **(a)** Representative chromatogram showing the peaks of DOPAC and HVA in the CSF of SMA patients. **(b,c)** DOPAC and HVA levels in SMA1 (n=26), SMA2 (n=17), SMA3 (n=12) patients as well as healthy control (HC) individuals (n=3). Data are shown as violin plots representing the median with interquartile range (IQR). Dots represent individual patients’ values.

We then investigated whether SMN induction with Nusinersen administration changes the CSF levels of dopamine metabolites. No significant differences in DOPAC and HVA levels were observed in SMA1 (n=17), SMA2 (n=15) and SMA3 (n=12) patients between T0 (baseline) and T302 (maintenance phase of Nusinersen therapy) (Figure 2a-c, Suppl. Table 3). The reduced number of patients at T302 was due to the unavailability of CSF samples, for reasons including missed follow-up visits or clinical circumstances preventing sample collection.

**Figure 2.**
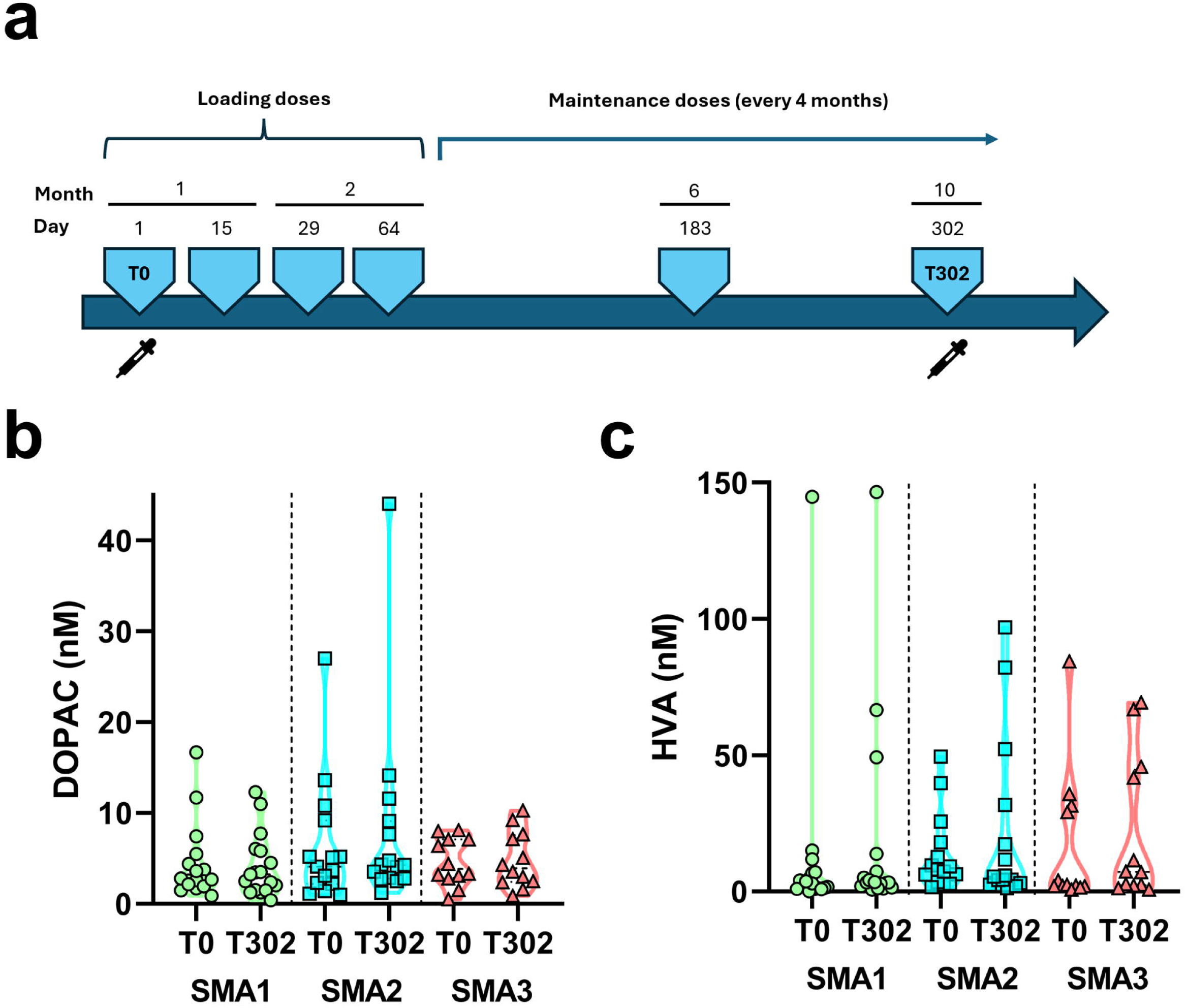
Effect of Nusinersen on CSF DOPAC and HVA levels in treated SMA patients. **(a)** Schematic representation of the timeline of intrathecal Nusinersen administration (12 mg/dose) and CSF collection in SMA patients. Sample collection was performed at baseline (T0) and at T302. **(b, c)** DOPAC and HVA levels in the CSF of SMA1 (n=17), SMA2 (n=15) and SMA3 (n=12) patients prior to treatment (T0) and at the time of the sixth (T302) injection of Nusinersen. Data are shown as violin plots representing the median with interquartile range (IQR). Dots represent individual patients’ values.

### Correlation of CSF dopamine-related metabolites with disease severity in naïve and Nusinersen-treated SMA patients

We next assessed the possible association between dopamine-related metabolites and clinical interventions reflecting disease severity, including nutritional and respiratory support. Since gastrostomy and tracheostomy were exclusively present in the most severely affected SMA1 patients of our cohort, subsequent analyses were restricted to this specific subgroup. Statistical analysis revealed that treatment-naïve SMA1 patients with gastrostomy (n=15) or tracheostomy (n=8) showed significantly lower CSF DOPAC levels than those without these interventions (n=11 and n=18, respectively) (*p*=0.009 and *p*=0.022, respectively; Figure 3a,c; Suppl. Table 4). SMA1 patients with gastrostomy were significantly older than those without this intervention (median age [IQR]: 4.0 [2.9-5.6] vs 0.8 [0.4-2.2] years, respectively; *p*=0.001), whereas patients with tracheostomy showed a non-significant age difference compared to those without (median age [IQR]: 4.3 [2.1-6.7] vs 2.6 [0.6-4.0] years; *p*=0.080). Notably, CSF DOPAC alterations were no longer significant after Nusinersen treatment, although a trend toward lower levels was maintained (Figure 3a,c; Suppl. Table 4). In contrast, no significant differences in CSF HVA levels were detected either at baseline or after treatment (Figure 3b,d; Suppl. Table 4).

**Figure 3.**
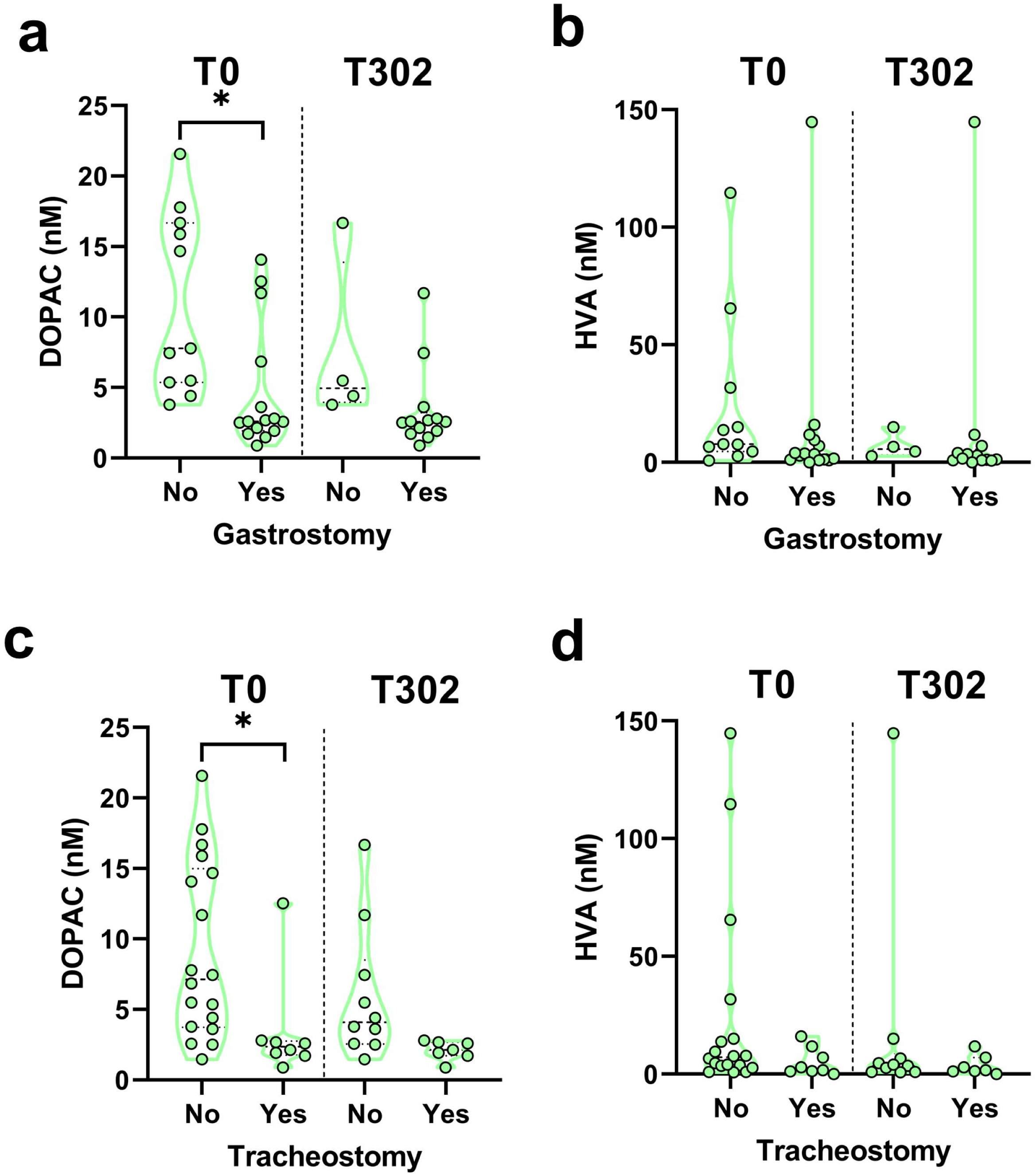
Basal and post-treatment CSF DOPAC and HVA levels in SMA1 patients stratified by gastrostomy or tracheostomy status. **(a,c)** DOPAC and **(b,d)** HVA levels in SMA1 patients requiring or not **(a,b)** gastrostomy (No, n=11; Yes, n=15) or **(c,d)** tracheostomy (No, n=18; Yes, n=8) at both baseline (T0) and at T302. Data are shown as violin plots representing the median with interquartile range (IQR). * *p*<0.05, compared with patients not requiring gastrostomy. P values derive from ANCOVA conducted on ln-transformed values (due to non-normally distributed residuals) with age as covariate. Dots represent individual patients’ values.

We then investigated the possible association between dopamine-related metabolite levels and SMN2 copy number across the entire SMA cohort. Moreover, we examined their relationship with the requirement for NIV; for this analysis, SMA1 and SMA2 patients were pooled, as just one patient in the SMA3 group required NIV. No significant differences in DOPAC or HVA concentrations emerged between SMA patients with fewer than three copies (n=26) and those with three or more copies (n=29) or between patients with (n=17) and without NIV (n=26) (Suppl. Table 5).

### Relationship of CSF dopamine-related metabolites with motor function in naïve and Nusinersen-treated SMA patients

We finally investigated the association of CSF DOPAC or HVA levels with motor function, assessed using CHOP-INTEND or HFMSE scores at baseline and after Nusinersen treatment. Non-parametric Spearman correlation analysis revealed no significant associations, except for a positive correlation between baseline DOPAC levels and CHOP-INTEND scores in SMA1 patients at T0 (n=26; *r*=0.531, *p*=0.005) (Suppl. Figure 1, Suppl. Table 6). However, after adjustment for age, this association did not retain statistical significance, although a clear positive trend persisted (*r*=0.532, *p*=0.062) (Suppl. Figure 1, Suppl. Table 6). Consistently, we also found a positive correlation between DOPAC levels and longitudinal changes achieved after treatment with Nusinersen, measured as ΔCHOP-INTEND (T302 – T0) (n=17; *r*=0.640, *p*=0.006) (Figure 4, Suppl. Table 7). Notably, this correlation remained statistically significant after correction for age (*r*=0.538, *p*=0.032), suggesting that higher baseline DOPAC levels are predictive of motor improvement after Nusinersen treatment.

**Figure 4.**
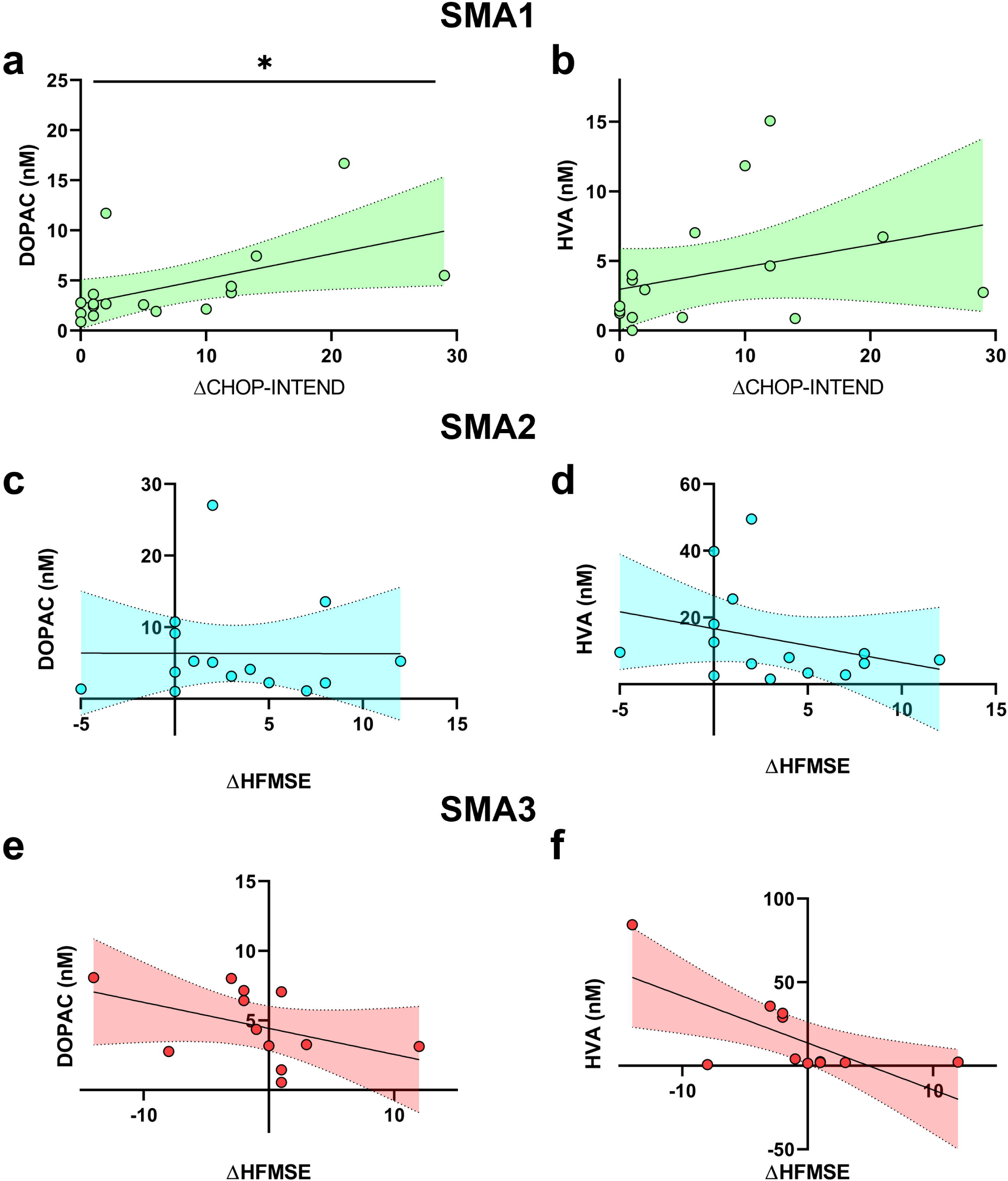
Partial correlations between CSF DOPAC or HVA levels at baseline and ΔCHOP-INTEND or ΔHFMSE in treated SMA patients. Correlation between **(a,c,e)** DOPAC or **(b,d,f)** HVA levels with **(a,b)** CHOP-INTEND or **(c-f)** HFMSE in **(a,b)** SMA1 (n=17), **(c,d)** SMA2 (n=15) and **(e,f)** SMA3 (n=12) patients. * *p*<0.05. P values derive from Spearman’s correlation, corrected by age.

## Discussion

The CSF levels of dopamine-related catabolites have been extensively investigated in Parkinson’s disease (PD) and atypical parkinsonism as biomarkers of nigrostriatal dopaminergic degeneration and disease progression^49–53^. A hallmark of PD is indeed the progressive loss of dopaminergic neurons in the substantia nigra, which is considered the main driver of motor symptoms^54^. Although fatigue, motor and postural impairment are also key clinical features of patients with SMA^2^, the involvement of central dopaminergic metabolism in this neurodegenerative disease has not been previously explored. In the present study, we addressed this issue by measuring the CSF levels of the two main dopamine catabolites, DOPAC and HVA, in SMA patients at baseline and after intrathecal treatment with Nusinersen.

We found no significant differences in baseline CSF DOPAC or HVA concentrations across SMA types, indicating the absence of a clear SMN-dependent alteration in dopamine-related catabolites. Although no significant association with age was detected, a downward trend in DOPAC and HVA levels from SMA1 to SMA3 was also observed. This trend is broadly consistent with the well-established age-related decline of these metabolites reported in healthy subjects^46–48,55^, given the progressive increase in median age across SMA types. However, the limited number of age-matched healthy controls precludes firm conclusions from these comparisons.

When analyzing clinical markers of disease severity in the most affected SMA1 subgroup, a distinct pattern emerged. Indeed, in treatment-naïve SMA1 patients, lower CSF DOPAC levels were associated with the presence of gastrostomy and tracheostomy, suggesting a connection between reduced dopamine turnover and more severe clinical status. Moreover, SMA1 patients requiring these supportive interventions tended to be older than those who did not, which aligns with a longer duration of the disease and a more advanced disease stage. Consistent with this evidence, SMA1 patients with lower baseline DOPAC levels demonstrated less improvement in motor function following Nusinersen treatment, as indicated by smaller changes in ΔCHOP-INTEND scores. Overall, these findings indicate that perturbed central dopaminergic metabolism is characteristic of advanced disease stages in SMA1, identifying patients with more severe motor impairment and reduced functional recovery.

The selective downregulation of CSF DOPAC levels observed in SMA1 infants requiring gastrostomy or tracheostomy may reflect the impact of these clinical interventions on mesocorticolimbic dopaminergic pathway activation in severe patients. In this regard, both human and animal studies have shown that chronic stress conditions are associated with reduced dopamine and dopamine-related metabolite levels in CSF and mesolimbic brain regions, respectively^56–61^. It is therefore plausible that prolonged disease burden exacerbated by invasive supportive interventions contributes to the dopaminergic system abnormalities observed in the most severe SMA1 patients.

We also acknowledge that the measured dopamine-related catabolite levels may have been affected by additional confounding factors. Indeed, both increased caloric intake and caloric restriction are known to influence central and peripheral catecholamine levels^62^. Accordingly, enteral feeding may have altered nutrient composition and energy balance, potentially impacting CNS dopaminergic metabolism in patients treated with gastrostomy. Further preclinical and human studies are thus warranted to elucidate the effect of these contributory factors.

Notably, in Nusinersen-treated patients, we found that SMN enhancement did not produce significant longitudinal changes in CSF DOPAC or HVA levels relative to baseline. Consistently, stratification of the entire cohort by SMN2 copy number or SMA subtype revealed no differences in CSF levels of the two dopamine-related metabolites, suggesting that CSF DOPAC and HVA levels are insensitive to SMN dosage-dependent effects in patients.

In contrast to clinical data, we previously reported that SMN deficiency in late-stage SMNΔ7 mice is associated with remarkable cerebral alterations of dopamine metabolism, including reduced AADC (aromatic L-amino acid decarboxylase) and increased MAO-A (monoamine oxidase A) expression, as well as enhanced TH (tyrosine hydroxylase) phosphorylation, ultimately converging toward increased cerebral dopamine content^28^. These findings are not necessarily discordant with the present clinical data, as CSF DOPAC reflects extracellular dopamine turnover and provides an integrated readout across multiple brain regions, rather than capturing tissue- or circuit-specific dopaminergic alterations^63^. Accordingly, SMN-driven changes in brain dopaminergic metabolism evident in experimental models may not be readily detectable from measurements of dopamine-related catabolites in patients. Discrepancies between preclinical and clinical settings, such as differences in disease stage, compensatory adaptations, and the burden of chronic stress, may further contribute to the varying neurochemical signatures observed. Overall, our findings suggest that CSF DOPAC levels mainly reflect disease stage and clinical severity, rather than the SMN-driven mechanisms that contribute to dopaminergic changes in SMA.

Supporting the functional relevance of dopaminergic dysregulation in SMA, a recent study demonstrated that loss of catecholaminergic synapses from spinal neurons contributes to neurological deficits in SMNΔ7^34^ mice and can be rescued by L-DOPA and benserazide treatment, supporting a reversible component of dopaminergic impairment in SMA. Notably, the number of TH-positive neurons in the substantia nigra pars compacta, ventral tegmental area^28^ and A11 hypothalamic region^34^ was comparable between SMNΔ7 and WT mice, suggesting that dopaminergic dysfunction in SMA arises from neurometabolic and synaptic alterations rather than dopaminergic neuron loss. Together with the present association between lower CSF DOPAC levels and greater disease severity observed in SMA patients, these findings support dopaminergic pathways as potential complementary therapeutic targets alongside SMN-enhancing strategies.

Alongside motor control, the dopamine system plays a crucial role in a wide range of cognitive processes, as well as mood and motivation^64–67^. Although neuropsychiatric outcomes were not assessed in the study protocol, dopaminergic dysregulation in advanced SMA may also mirror the presence of specific neurobehavioral disturbances, which are relatively common in severe SMA patients^68^.

Lower CSF concentrations of dopamine-related catabolites have been associated with greater motor dysfunction^52,53^ and loss of dopaminergic terminals in PD^51,52,69^. Although the dopaminergic pathways involved in PD and SMA pathogenesis are different (nigrostriatal^54^ versus A11 hypothalamic projections^34^), the CSF level of dopamine products may represent useful markers of dopaminergic dysmetabolism and motor dysfunction across neurodegenerative disorders.

Despite its novelty, our exploratory study has some limitations, including the relatively small sample size and the exclusive analysis of CSF, which reflects a composite signal from multiple CNS regions^70–73^. Although we adopted a standardized CSF collection procedure^28^, these factors may have partially contributed to variability in metabolite levels.

In conclusion, this work provides the first evidence that reduced central DOPAC levels are associated with greater disease severity and poorer motor response in SMA1 patients, suggesting a possible role for dopaminergic metabolism dysfunction in the most severe forms of SMA. Given the growing recognition of SMA as a multisystem disease^74^, the identification of dopaminergic neurotransmission as a potentially involved system may open new possibilities for adjunctive therapeutic strategies that integrate SMN-targeted approaches with dopaminergic modulation, especially in patients with severe disease.

## Data availability

The data that support the findings of this study are available from the corresponding author upon reasonable request.

## Authors contributions

T.N.: Formal analysis, Visualization, Writing – review & editing; B.R.: Formal analysis, Visualization, Writing – original draft, Writing – review & editing; V.B.: Investigation; A.D.A.: Investigation, Data curation, Resources, Writing – review & editing; A.I.: Formal analysis, Writing – original draft, Writing – review & editing; A.L.: Investigation, Data curation; M.C.: Investigation; C.P.: Investigation, Data curation, Resources; C.B.: Investigation, Data curation, Resources, Writing – review & editing; E.M.V.: Writing – review & editing; M.F.: Writing – review & editing; E.B.: Investigation, Data curation, Resources, Writing – review & editing; F.E.: Writing – review & editing, Supervision; A.U.: Conceptualization, Writing – review & editing, Supervision.

