## Supplementary information for "Disruption of central dopamine metabolism in infants with severe spinal muscular atrophy"

**Dysregulation of central dopamine metabolism in children with severe**

**spinal muscular atrophy**

**Supplementary Table 1. CSF DOPAC and HVA levels in treatment-naïve SMA patients.**

|  | **Controls**  **(n=3)** | **SMA1**  **(n=26)** | **SMA2**  **(n=17)** | **SMA3**  **(n=12)** | ***p* value^1^** | ***p* value^2^** |
| --- | --- | --- | --- | --- | --- | --- |
| **DOPAC (nM)** | 5.5 (3.5-13.3) | 4.9 (0.9-21.6) | 5.1 (1.0-27.0) | 3.8 (0.5-8.1) | 0.755 | 0.981 |
| **HVA (nM)** | 9.5 (5.6-16.0) | 5.7 (1.6-14.1) | 8.0 (4.8-19.5) | 2.3 (1.9-30.9) | 0.347 | 0.218 |

Data are expressed as median (interquartile range, IQR). ^1^Group comparisons were performed using a non-parametric Kruskal-Wallis test. ^2^ANCOVA was conducted on ln-transformed values (due to non-normally distributed residuals) with age as a covariate. The number (n) of patients is indicated. Abbreviations: 3,4-dihydroxyphenylacetic acid (DOPAC), homovanillic acid (HVA).

**Supplementary Table 2. Correlation between CSF DOPAC or HVA levels and age in treatment-naïve SMA patients.**

| **Parameter** | **Patients** | **DOPAC (nM)** | **HVA (nM)** |
| --- | --- | --- | --- |
| **Age (years)** | **SMA1-2-3 (n=55)** | *r=*-0.196 | *r=*0.032 |
|  |  | *p*=0.152 | *p*=0.818 |
|  | **SMA1 (n=26)** | ***r=*-0.451** | *r=*-0.198 |
|  |  | ***p*=0.021** | *p*=0.332 |
|  | **SMA2 (n=17)** | *r=*0.040 | *r=*0.256 |
|  |  | *p*=0.877 | *p*=0.321 |
|  | **SMA3 (n=12)** | *r=*-0.042 | *r=*0.098 |
|  |  | *p*=0.897 | *p*=0.762 |

Non-parametric Spearman’s rho coefficients (*r*) and related *p* values (*p*) are indicated. Significant *p* values are shown in bold. The number (n) of patients is indicated. Abbreviations: 3,4-dihydroxyphenylacetic acid (DOPAC), homovanillic acid (HVA), total cohort (tot).

**Supplementary Table 3. Longitudinal changes in CSF DOPAC and HVA levels in Nusinersen-treated SMA patients between T0 and T302.**

| **Metabolite** | **SMA1 (n=17)** | **SMA2 (n=15)** | **SMA3 (n=12)** |
| --- | --- | --- | --- |
|  | **T0 vs T302** | **T0 vs T302** | **T0 vs T302** |
| **DOPAC (nM)** | 2.6 vs 3.2 (*p* = 0.981) | 4.1 vs 4.3 (*p* = 0.173) | 3.8 vs 3.9 (*p* = 0.583) |
| **HVA (nM)** | 2.9 vs 3.3 (*p* = 0.687) | 8.0 vs 5.4 (*p* = 0.865) | 2.3 vs 7.2 (*p* = 0.209) |

Data are expressed as median. *p* values derive from non-parametric Wilcoxon test. The number (n) of patients is indicated. Abbreviation: 3,4-dihydroxyphenylacetic acid (DOPAC), homovanillic acid (HVA).

**Supplementary Table 4. Basal and post-treatment CSF DOPAC and HVA levels in SMA1 patients, stratified by the presence of gastrostomy or tracheostomy.**

|  | **T0** | | | **T302** | | |
| --- | --- | --- | --- | --- | --- | --- |
| **Metabolite** | **Gastrostomy** | | ***p value*** | **Gastrostomy** | | ***p value*** |
|  | **No (n=11)** | **Yes (n=15)** |  | **No (n=4)** | **Yes (n=13)** |  |
| **DOPAC (nM)** | 7.8 (5.4-16.7) | 2.6 (1.9-6.8) | **0.009** | 4.0 (2.0-5.5) | 2.5 (1.4-6.9) | 0.421 |
| **HVA (nM)** | 7.8 (4.6-31.8) | 3.6 (1.2-9.5) | 0.084 | 4.0 (1-6-111.2) | 3.3 (1.7-10.6) | 0.57 |
| **Metabolite** | **Tracheostomy** | | ***p value*** | **Tracheostomy** | | ***p value*** |
|  | **No (n=18)** | **Yes (n=8)** |  | **No (n=10)** | **Yes (n=7)** |  |
| **DOPAC (nM)** | 7.1 (3.7-15) | 2.4 (1.8-2.8) | **0.022** | 3.3 (1.5-7.1) | 2.5 (1.2-6) | 0.903 |
| **HVA (nM)** | 7.2 (3.4-19.2) | 2.3 (1.3-10.6) | 0.199 | 3.8 (1.3-22.1) | 2.3 (2-13.9) | 0.814 |

Data are expressed as median (interquartile range, IQR). *p* values derive from ANCOVA conducted on ln-transformed values with age as covariate. The number (n) of patients is indicated. Abbreviation: 3,4-dihydroxyphenylacetic acid (DOPAC); homovanillic acid (HVA).

**Supplementary Table 5.** **Basal and post-treatment CSF DOPAC and HVA levels in SMA patients, stratified by SMN2 copy number or the presence of non-invasive ventilation.**

|  | **T0** | | | **T302** | | |
| --- | --- | --- | --- | --- | --- | --- |
| **Metabolite** | **SMN2 cn  (SMA 1-2-3)** | | ***p value*** | **SMN2 cn  (SMA 1-2-3)** | | ***p value*** |
|  | **< 3 (n=26)** | **≥ 3 (n=29)** |  | **< 3 (n=18)** | **≥ 3 (n=26)** |  |
| **DOPAC (nM)** | 3.7 (2.4-12.4) | 5.1 (2.9-8.0) | 0.249 | 3.3 (1.9-5.9) | 4.2 (2.6-8.0) | 0.188 |
| **HVA (nM)** | 3.9 (1.6-12.3) | 7.7 (2.7-27.4) | 0.281 | 3.3 (1.8-7.3) | 6.4 (2.6-47.3) | 0.422 |
| **Metabolite** | **NIV (SMA1-2)** | | ***p value*** | **NIV (SMA1-2)** | | ***p value*** |
|  | **No (n=26)** | **Yes (n=17)** |  | **No (n=19)** | **Yes (n=13)** |  |
| **DOPAC (nM)** | 4.3 (2.2-16.3) | 5.2 (2.5-12.9) | 0.334 | 3.6 (2.0-7.7) | 3.2 (2.4-6.7) | 0.813 |
| **HVA (nM)** | 6.6 (2.5-9.9) | 9.6 (3.7-28.7) | 0.151 | 3.3 (2.0-13.8) | 4.3 (3.8-24.5) | 0.263 |

Data are expressed as median (interquartile range, IQR). *p* values derive from ANCOVA conducted on ln-transformed values with age as covariate. The number (n) of patients is indicated. Abbreviation: 3,4-dihydroxyphenylacetic acid (DOPAC), homovanillic acid (HVA), non-invasive ventilation (NIV), SMN2 copy number (SMN2 cn).

**Supplementary Table 6. Correlation between CSF DOPAC or HVA levels and functional motor scales of SMA patients before and after Nusinersen treatment.**

| **Functional motor scale** | **Patients** | **Therapy phase** | **DOPAC (nM)** | **HVA (nM)** |
| --- | --- | --- | --- | --- |
| **CHOP-INTEND** | **SMA1** | **T0** | ***r=*0.531** | *r=*0.297 |
|  |  |  | ***p=0.005^a^*** | *p=0.141* |
|  |  |  | n=26 | n=26 |
|  |  | **T302** | *r=*0.264 | *r=*0.040 |
|  |  |  | *p=0.306* | *p=0.877* |
|  |  |  | n=17 | n=17 |
| **HFMSE** | **SMA2** | **T0** | *r=*0.080 | *r=*-0.177 |
|  |  |  | *p=0.758* | *p=0.496* |
|  |  |  | n=17 | n=17 |
|  |  | **T302** | *r=*-0.034 | *r=*-0.500 |
|  |  |  | *p=0.904* | *p=0.058* |
|  |  |  | n=15 | n=15 |
|  | **SMA3** | **T0** | *r=*0.120 | *r=*0.380 |
|  |  |  | *p=0.715* | *p=0.223* |
|  |  |  | n=12 | n=12 |
|  |  | **T302** | *r=*0.000 | *r=*0.042 |
|  |  |  | *p=>0.999* | *p=0.897* |
|  |  |  | n=12 | n=12 |

Non-parametric Spearman’s rho coefficients (r) and related *p* values (*p*) are indicated. Significant *p* values are shown in bold. ^a^ The *p* value does not survive after correction for age (r =0.532, *p* =0.062). Abbreviations: Children’s Hospital of Philadelphia Infant Test of Neuromuscular Disorders (CHOP-INTEND), Hammersmith Functional Motor Scale Expanded (HFMSE), 3,4-dihydroxyphenylacetic acid (DOPAC), homovanillic acid (HVA).

**Supplementary Table 7. Correlation between baseline CSF DOPAC or HVA levels and changes in functional motor scales in SMA patients after treatment with Nusinersen.**

| **Functional motor scale** | **Patients** | **DOPAC (nM)** | **HVA (nM)** |
| --- | --- | --- | --- |
| **ΔCHOP-INTEND** | **SMA1 (n=17)** | ***r=*0.640** | *r=0.350* |
|  |  | ***p*=0.006^a^** | *p*=0.169 |
| **ΔHFMSE** | **SMA2 (n=15)** | *r=0.054* | *r=-0.394* |
|  |  | *p*=0.848 | *p*=0.147 |
|  | **SMA3 (n=12)** | *r=-0.536* | *r=-0.434* |
|  |  | *p*=0.072 | *p*=0.159 |

Non-parametric Spearman’s rho coefficients (*r*) and related *p* values (*p*) are indicated. Significant *p* values are shown in bold. ^a^ The *p* value survives after correction for age (*r=*0.538*, p*=0.032). Abbreviations: Delta (Δ) [T302-T0], Children’s Hospital of Philadelphia Infant Test of Neuromuscular Disorders (CHOP-INTEND), Hammersmith Functional Motor Scale Expanded (HFMSE), 3,4-dihydroxyphenylacetic acid (DOPAC), homovanillic acid (HVA).

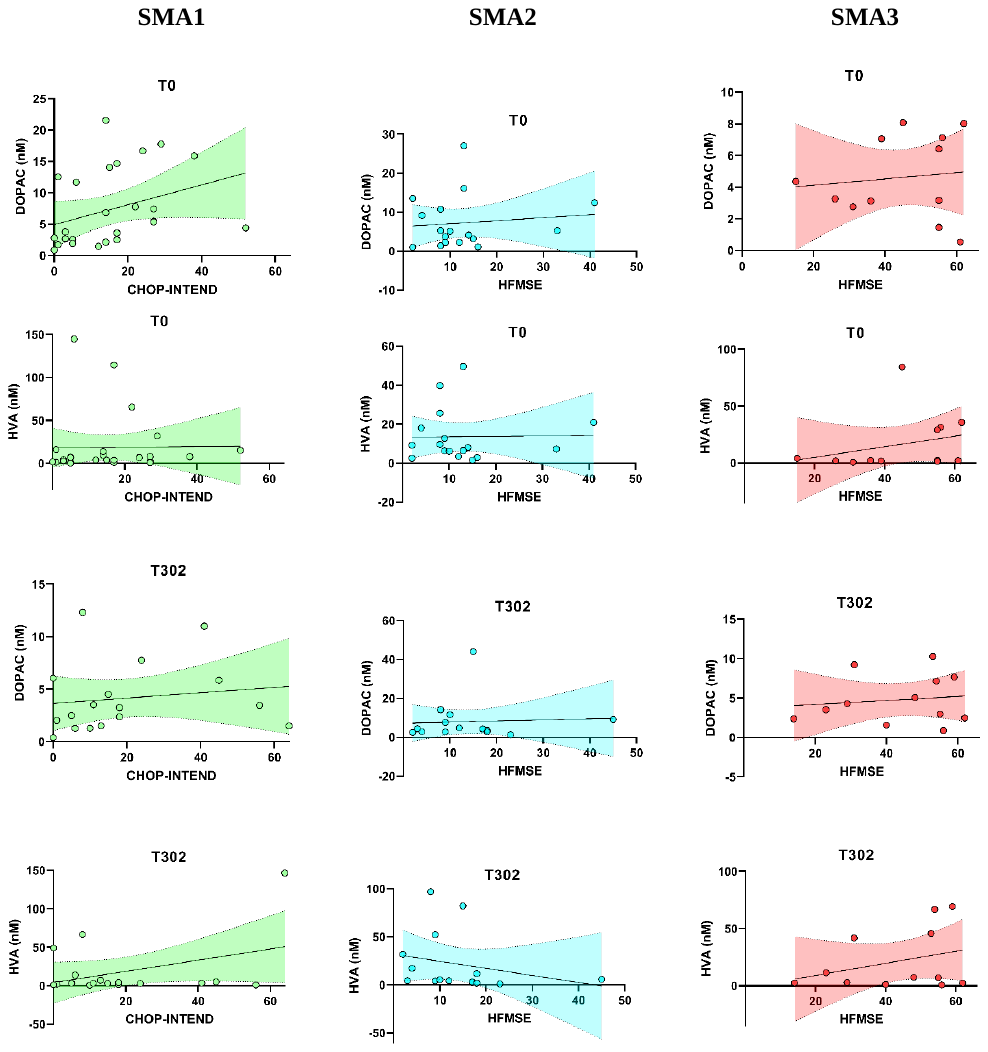

**Supplementary Figure 1. Age-adjusted partial correlations between baseline CSF DOPAC or HVA levels and functional motor scales in naïve and Nusinersen-treated SMA patients.** Partial correlation analyses (corrected for age) between CSF levels of 3,4-dihydroxyphenylacetic acid (DOPAC) or homovanillic acid (HVA) with motor functions are shown for SMA1, SMA2 and SMA3 patients at baseline (T0) and after Nusinersen treatment (T302). Motor function was evaluated using CHOP-INTEND in SMA1 patients and HFMSE in SMA2 and SMA3 patients.
